# Investigating Inflammation and Tissue Remodeling in ILD with [^11^C]NES and [^68^Ga]Ga-FAPI-46 PET Imaging

**DOI:** 10.1101/2025.06.18.25329561

**Authors:** Olivia Wegrzyniak, Emil Ekbom, Jens Ellingsen, Olof Eriksson, Mark Lubberink, Andrei Malinovschi, Jonathan Sigfridsson, Irina Velikyan, Viola Wilson, Össur Emilsson, Gunnar Antoni

**Affiliations:** Science for Life Laboratory, Department of Medicinal Chemistry, Uppsala University, Uppsala, Sweden; Department of Medical Sciences: Respiratory-, allergy and sleep research, Uppsala University, Uppsala, Sweden; Integrative Physiology, Department of Medical Cell Biology, Uppsala University, Sweden; Department of Surgical Sciences, Uppsala University, Uppsala, Sweden; Clinical Physiology, Department of Medical Sciences, Uppsala University; PET-center, Uppsala University Hospital, Uppsala, Sweden; Department of Medicinal Chemistry, Uppsala University, Uppsala, Sweden

**Keywords:** Lung fibrosis, inflammation, neutrophil, FAP, PET

## Abstract

Interstitial lung diseases (ILDs) encompass a large number of conditions that affect the lungs, characterized by varying degrees of inflammation and fibrosis. Accurate diagnosis and monitoring of ILDs remain challenging due to the complex nature of ILDs and the limitations of conventional diagnostic methods such as high-resolution computed tomography and pulmonary function tests.

**Methods:** Fourteen patients with different ILDs and four healthy controls underwent positron emission tomography (PET) imaging using [^11^C]NES followed by [^68^Ga]Ga-FAPI-46, to visualize and quantify neutrophil elastase and fibroblast activated protein in the lungs, respectively. On the same day of the PET examination, blood samples were collected from the participant to measure various immune biomarkers in order to compare between PET results, serum biomarker levels, and clinical observations.

**Results:** Uptake of both radiotracers were higher in the lungs of ILD patients compared to healthy controls, with particularly strong uptake in regions showing CT features of fibrosis. [^68^Ga]Ga-FAPI-46 generally exhibited higher uptake than [^11^C]NES in these fibrotic areas. Despite the elevation of several immune biomarkers compared to healthy controls, no clear correlation was found between these biomarkers and PET imaging results.

**Conclusion:** These preliminary results support the potential of [^11^C]NES and [^68^Ga]Ga-FAPI-46 PET/CT imaging as promising non-invasive methods for assessing neutrophil-mediated inflammation and tissue remodeling activity in the lungs of ILD patients.

## INTRODUCTION

Interstitial lung diseases (ILDs) are a large and heterogeneous group of several disorders affecting the lungs with varying degrees of inflammation and fibrosis which can lead to respiratory failure^1^.

Monitoring ILD disease activity remains challenging. Conventional diagnostic methods, such as high-resolution computed tomography (HRCT), detect specific morphologic pathological patterns and help establish the diagnosis of ILDs. Pulmonary function tests (PFTs) measure the decrease in lung function.^2,3^ However, these approaches do not directly quantify tissue remodeling activity and only assess the cumulative result of tissue damage, which can lead to delayed diagnosis. Additionally, histopathological examination of lung biopsies, while informative, is an invasive technique with its associated limitations (complexity, cost, postoperative complications…)^4,5^.

There is a critical need for non-invasive techniques that can accurately assess and monitor fibrosis-driven as compared with inflammation-driven disease activity in ILD patients. Such methodologies would be highly valuable for the early assessment of disease severity, facilitate the selection of suitable treatment (anti-inflammation or anti-fibrotic), and monitor treatment response. This could also contribute to the development of anti-fibrotic therapies, addressing the current limitations in treatment options.

Positron emission tomography (PET) imaging with radioligands targeting fibroblast activation protein (FAP) has shown promise in detecting activated fibroblasts in ILD and in monitoring fibrotic changes post-treatment^6–9^. Similarly, [^11^C]NES, which targets neutrophil elastase (NE), has proven effective in assessing neutrophil-driven inflammation in COVID-19 patients with severe lung inflammation^10^ and with long-term pulmonary complications^[manuscript post– covid].^

This study hypothesizes that PET imaging with [^11^C]NES and [^68^Ga]Ga-FAPI-46 can effectively quantify inflammation through neutrophil activity and tissue remodeling through fibroblast activation, respectively, in patients with ILD of varying severity and etiology.

## MATERIALS AND METHODS

### Subjects

In this pilot, 14 patients previously diagnosed with ILD and 4 healthy controls were recruited at the lung clinic of the University Hospital of Uppsala (Sweden). Patients receiving antifibrotic or immunosuppressive therapy were excluded, with the exception of those receiving prednisolone at doses up to 10mg/day. The participants were consecutively examined with two PET tracer molecules: [^11^C]NES and [^68^Ga]Ga-FAPI-46. The PET/CT examinations were performed at the PET Center of the University Hospital of Uppsala between January 2022 and May 2024. The blood of each participant was taken the day of the scans before injection, to be later analyzed for immune cell counts and Olink analyses. Six patients with ILD also underwent a biopsy, within months after the PET examination, which was routinely analyzed by a pathologist from Uppsala University.

### PET/CT examination

[^11^C]NES and [^68^Ga]Ga-FAPI-46 were produced as previously described [post-covid manuscript].

PET/CT examinations were conducted using a Discovery MI-5 PET/CT scanner (GE Healthcare) using a 25 cm, axial field of view. The participants underwent three consecutive scans with different radiotracers.

First, the participants were injected with a controlled bolus injection of [^11^C]NES (mean injected dose: 4.76±0.27MBq/kg, approximately 2-4µg/mL of [^11^C]NES). A PET scan was initiated simultaneously with the tracer injection, with a duration of 30min. Finally, after at least 2 hours following the [^11^C]NES injection, a 60min dynamic PET scan commenced simultaneously with the administration of [^68^Ga]Ga-FAPI-46 (mean injected dose: 2.39±0.52MBq/kg, approximately 2-4µg of [^68^Ga]Ga-FAPI-46). Whole-body PET/CT examinations of 3min were performed after the dynamic [^11^C]NES and [^68^Ga]Ga-FAPI-46 PET scans.

Dynamic and static whole-body PET/CT images were reconstructed as previously described^10^. Further details regarding the PET/CT examination can be found in the supplementary data.

### PET/CT analysis

Volumes of interest (VOIs) were manually delineated over the lungs (Fig.S1). Whole-lung VOIs were defined on pulmonary parenchyma in the left and right lung. The inflammation volume (I.V.) and remodeling volume (R.V.) were identified as the volume with visible uptake in the left and right lung with a standardized uptake value (SUV) ranging from 0.75 (corresponding to background lung uptake) to 3, on the final frame of the dynamic PET scans (1500-1800s and 3000-3600s for [^11^C]NES and [^68^Ga]Ga-FAPI-46). The basal part of the lung and the signal observed around the lobar arteries and veins were excluded from I.V. and R.V.. Additionally, VOIs were defined in the apical segment (where tracer uptake was typically lower) and in the basal part of the lung.

VOIs were also delineated on the dynamic images for the aorta, liver, spleen, muscle, and bone marrow, and time-activity curves generated for all organs. VOIs of different organs of interest were also delineated on the static whole-body scans. The analysis was performed using PMOD software (PMOD Technologies LLC), and the SUV_mean_, SUV_max_, and SUV_tot_ (for whole-lung VOIs) were obtained for these regions. SUV_tot_ in each lung was calculated by multiplying the SUV_mean_ of the lung ROI by the total whole lung volume.

The representative chest PET/CT images shown here correspond to the last frame of the scans.

### Proximity Extension analysis (PEA)

Two proximity extension assays (PEA) were conducted: one using the Olink® Target 96 Inflammation panel and the other using the Olink® Target 48 Cytokine panel. For the first assay, blood samples from healthy controls from a previous study (n=2) and from patients ILD01 to ILD11 (n=11) were analyzed. For the second assay, blood samples from four healthy controls from this study, the two healthy controls from the previous study (n=6), and from all ILD patients except ILD12 (n=13) were included.

Blood samples were collected in EDTA tubes on the day of the PET scans, except for the samples from the two healthy controls from the previous study. Plasma was isolated by centrifuging the blood samples in EDTA tubes at 4500 rpm for 5 minutes at 4°C. The plasma was then extracted into cryotubes and stored at -80°C until the PEA analyses.

For the assays, 25 µL of plasma was placed at the bottom of each V-shaped well in a 96-well plate. Each sample from healthy controls and ILD patients was measured in four replicates for the Olink® Target 96 Inflammation panel and in duplicate for the Olink® Target 48 Cytokine panel. Plasma protein concentrations were analyzed using the two Olink® panels by the SciLifeLab Unit of Affinity Proteomics in Uppsala, Sweden. Only samples that passed quality control tests were reported. The results were expressed as normalized protein expression (NPX) on a log2 scale for the Olink® Target 96 Inflammation panel and as pg/mL values for the Olink® Target 48 Cytokine panel.

### Statistical analysis

Given the small study cohort, p-values are exploratory. The lung SUV uptake comparison was made using Kruskal Wallis and uncorrected Dunn’s post-hoc tests. Spearman’s correlation was used for associations between tracer uptake, clinical parameters, and serum biomarkers. These analyses were performed using GraphPad Prism 10.2.3. Olink panel data were analyzed using Olink’s Stat Analyzer tool. The protein levels presented in the heatmap were normalized into Z-scores before analysis. A significance level of 0.05 was used for all tests.

## RESULTS

The study recruited fourteen ILD patients and four controls, and their characteristics are detailed in Table 1 and supplemental Table 1 and 2. Two patients with ILD and one control did not complete the full study procedure. Patient ILD02 stopped after the [^11^C]NES dynamic chest PET/CT due to claustrophobia, and control HC04 stopped during the [^68^Ga]Ga-FAPI-46 dynamic chest PET/CT. Additionally, data for patient ILD13’s [^11^C]NES scan are missing due to a technical issue occurring during tracer labeling. Of the fourteen ILD patients, six had a histopathologically confirmed diagnosis within two months of the PET scans. The observations from the biopsies are presented in Table S2.

**Table 1.** Clinical characteristics of the patients.

| Variable |  |  |
| --- | --- | --- |
| Age (Years)* |  | 73.5(68.25;78.25) |
| Sex | Male | 11(79%) |
|  | Female | 3(21%) |
| BMI (kg/m2)* |  | 28(25.75;30.75) |
| Smoking status | Former smokers | 5(36%) |
|  | Never smoked | 9(64%) |
| Co-morbid diseases (n (%)) | Hypertension | 6(43%) |
|  | Diabetes | 4(29%) |
|  | Cancer | 1(7%) |
|  | Pulmonary hypertension | 3(21%) |
|  | Rheumatoid Arthritis | 3(21%) |
|  | Cirrhosis | 1(7%) |
| Blood analysis* | Hemoglobin (g/L) | 140.5(125.5;156) |
|  | Sedimentation rate (mm/h) | 16(7.75;38.75) |
|  | C-reactive protein (mg/dL) | 2.1(1.28;4.5) |
|  | Neutrophils (cells.10<sup>9</sup>/L) | 4.45(3.55;5) |
|  | Lymphocytes (cells.10<sup>9</sup>/L) | 78.41(63.08; 91.44) |
| Pulmonary function test<br>*(n=13) | FVC(%) | 2.87(2.16; 4.12) |
|  | TLC(%) | 69.5(61.75;77.25) |
|  | DLCO(%) | 52.5(43.5;64.5) |
| 6MWD(m)(n=12)* |  | 452.5(302;550.3) |

\*Data are presented as median (IQR25; IQR75). BMI, body mass index; DLCO, diffusing capacity for carbon monoxide; FVC, forced vital capacity; TLC, total lung capacity; 6MWD, 6min walk distance.

### Tracer uptake in the fibrotic lungs

In the lungs of ILD patients, high uptake of both tracers co-localized with areas showing CT signs of inflammation and fibrosis, while uptake was low in the normal parts of the lung (apical region) (Fig. 1, S2,S3 and Table 2,3). In contrast, only background uptake was observed in healthy lungs (Fig2, S2). Both ILD patients and healthy individuals showed tracer accumulation in the dorsal-basal part of the lungs. Interestingly, this basal-dorsal uptake was reduced when scanning a patient in the prone position (Fig S4).

**Figure 1.**
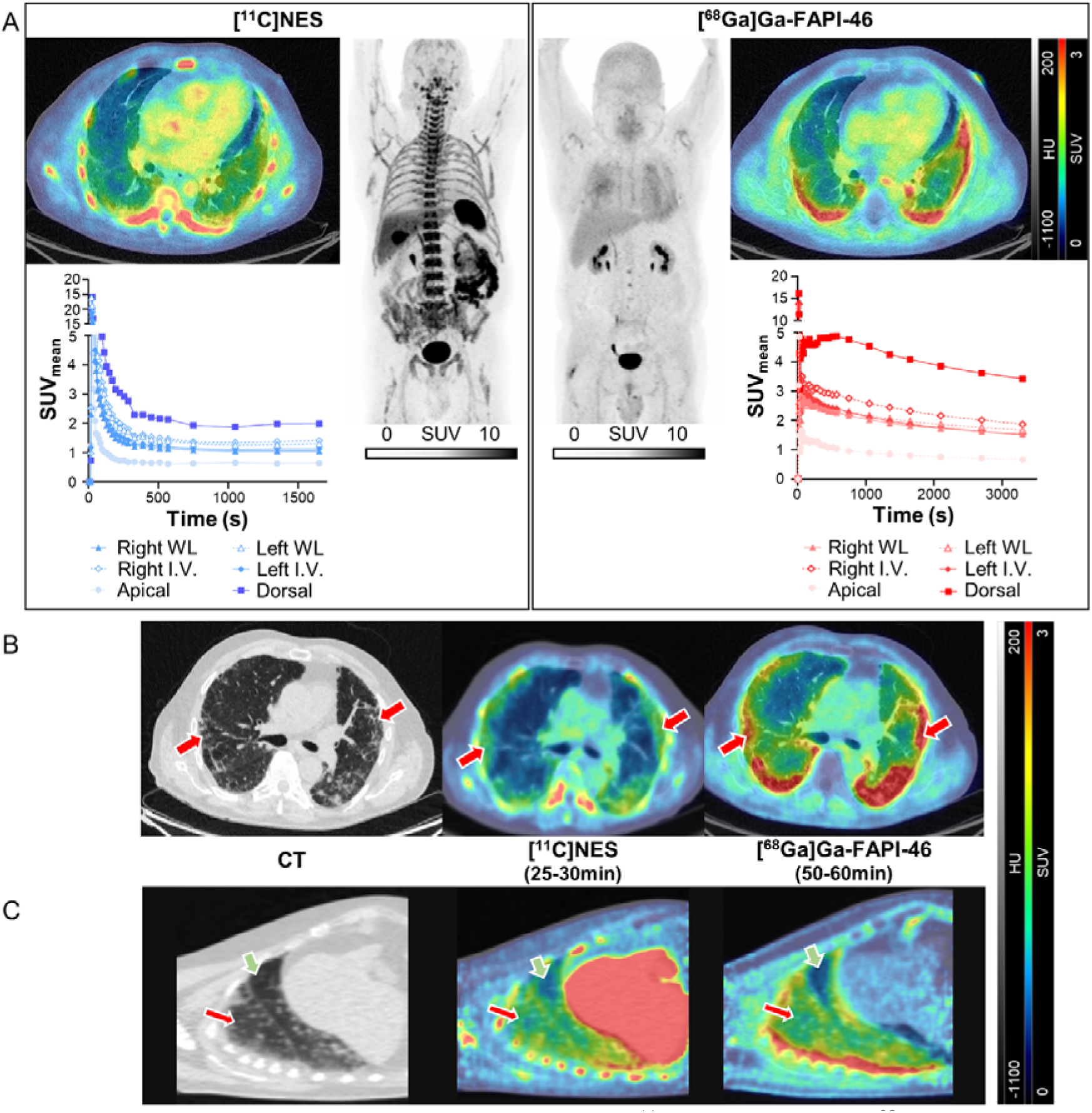
Tracers’ uptakes in fibrotic lung tissue. **(A)** [^11^C]NES (left) and [^68^Ga]Ga-FAPI-46-PET/CT (right) uptake in a patient with IPF (ILD14): full-body maximum intensity projection, axial PET/CT, and corresponding time–activity curve showing tracers’ lung uptake over time. **(B-C)** Axial and sagittal CT and PET/CT images of the tracers’ positive uptakes in lungs area with CT signs of fibrosis and inflammation (red arrows), but not in non-fibrotic area (green arrows), in a patient with IPF with PPF (ILD12) **(B)**, and another one with NSIP with PPF (ILD01) **(C)**. HU, Hounsfield unit; I.V.,inflammation volume; R.V., remodeling volume; SUV,standardized uptake value; WL,whole lung.

**Table 2.** [^11^C]NES PET/CT derived measures in all subjects.

| | [$^{11}\text{C}$]NES | | | | | | | | |
| --- | --- | --- | --- | --- | --- | --- | --- | --- | --- |
|  | WL Left |  |  | WL Right |  |  | I.V.* |  | V (cm<sup>3</sup>) |
|  | SUV<sub>mean</sub> | SUV<sub>tot</sub> | SUV<sub>max</sub> | SUV<sub>mean</sub> | SUV<sub>tot</sub> | SUV<sub>max</sub> | SUV<sub>mean</sub> | SUV<sub>max</sub> |  |
| ILD01 | 1.79 | 856.0 | 3.86 | 1.49 | 891.1 | 3.85 | 1.58 | 3.18 | 313.1 |
| ILD02 | 2.45 | 1540.9 | 5.25 | 0.84 | 1336.1 | 2.81 | 2.07 | 5.25 | 503.8 |
| ILD03 | 2.22 | 2147.4 | 6.06 | 1.84 | 2290.0 | 5.62 | 2.14 | 5.05 | 777.5 |
| ILD04 | 0.88 | 697.3 | 2.02 | 0.73 | 972.2 | 2.10 | 1.11 | 2.02 | 91.1 |
| ILD05 | 1.28 | 1303.5 | 2.54 | 1.10 | 1146.7 | 2.51 | 1.47 | 2.54 | 530.0 |
| ILD06 | 1.34 | 1215.6 | 3.77 | 1.18 | 1364.6 | 3.70 | 1.53 | 2.91 | 352.1 |
| ILD07 | 1.17 | 1083.0 | 2.89 | 1.06 | 1302.8 | 2.74 | 1.38 | 2.15 | 158.6 |
| ILD08 | 1.62 | 746.9 | 3.63 | 1.15 | 944.4 | 2.46 | 1.42 | 3.12 | 1042.4 |
| ILD09 | 1.67 | 810.9 | 3.54 | 1.48 | 972.3 | 4.19 | 1.71 | 3.54 | 367.3 |
| ILD10 | 1.18 | 1106.0 | 3.54 | 1.01 | 1069.3 | 2.79 | 1.27 | 2.36 | 362.6 |
| ILD11 | 1.26 | 908.3 | 2.98 | 0.95 | 1174.7 | 2.48 | 1.38 | 2.45 | 180.1 |
| ILD12 | 1.01 | 909.3 | 3.38 | 0.80 | 1176.0 | 2.49 | 1.47 | 2.73 | 641.3 |
| ILD13 | n.a | n.a | n.a | n.a | n.a | n.a | n.a | n.a | n.a |
| ILD14 | 1.13 | 822.5 | 2.96 | 1.05 | 919.8 | 2.41 | 1.36 | 2.17 | 202.5 |
| HC01 | 0.49 | 657.5 | 3.72 | 0.51 | 848.5 | 2.20 | n.a | n.a | n.a |
| HC02 | 0.79 | 646.6 | 2.55 | 0.72 | 783.0 | 2.75 | n.a | n.a | n.a |
| HC03 | 0.72 | 760.7 | 4.45 | 0.63 | 831.3 | 4.13 | n.a | n.a | n.a |
| HC04 | 0.74 | 524.1 | 2.39 | 0.70 | 690.3 | 3.02 | n.a | n.a | n.a |

\*The inflammation volumes (I.V.) do not include the basal posterior part of the lung with dependent atelectasis while the whole lung (WL) volume does. I.V. is lung volume with SUV>0.75 for [ $^{11}\text{C}$ ]NES.

**Table 3.** [^68^Ga]Ga-FAPI-46 PET/CT derived measures in all subjects.

|  | [<sup>68</sup>Ga]Ga-FAPI-46 |  |  |  |  |  |  |  |  |
| --- | --- | --- | --- | --- | --- | --- | --- | --- | --- |
|  | WL Left |  |  | WL Right |  |  | R.V.* |  |  |
|  | SUV<sub>mean</sub> | SUV<sub>tot</sub> | SUV<sub>max</sub> | SUV<sub>mean</sub> | SUV<sub>tot</sub> | SUV<sub>max</sub> | SUV<sub>mean</sub> | SUV<sub>max</sub> | V (cm<sup>3</sup>) |
| **ILD01** | 1.55 | 1064.5 | 3.79 | 1.60 | 1338.3 | 4.97 | 1.76 | 3.58 | 454.3 |
| **ILD02** | n.a | n.a | n.a | n.a | n.a | n.a | n.a | n.a | n.a |
| **ILD03** | 1.51 | 1173.6 | 7.06 | 1.58 | 1636.3 | 6.43 | 1.91 | 6.55 | 740.6 |
| **ILD04** | 1.16 | 977.9 | 3.28 | 0.89 | 1278.2 | 3.24 | 1.40 | 2.81 | 193.4 |
| **ILD05** | 1.37 | 1556.9 | 3.61 | 1.61 | 1842.8 | 4.20 | 1.73 | 3.91 | 870.7 |
| **ILD06** | 2.32 | 2243.2 | 7.92 | 1.83 | 2397.1 | 9.65 | 2.59 | 5.16 | 307.7 |
| **ILD07** | 1.29 | 1510.3 | 3.17 | 1.28 | 1787.5 | 3.16 | 1.51 | 2.90 | 624.1 |
| **ILD08** | 3.31 | 1979.1 | 7.16 | 1.76 | 2081.8 | 4.45 | 2.66 | 7.16 | 1025.7 |
| **ILD09** | 1.68 | 998.0 | 3.73 | 1.51 | 1163.8 | 3.83 | 1.80 | 3.37 | 421.6 |
| **ILD10** | 0.90 | 1161.1 | 2.91 | 0.91 | 1427.9 | 3.81 | 1.39 | 2.86 | 330.1 |
| **ILD11** | 1.79 | 1566.6 | 4.46 | 1.33 | 1808.3 | 4.26 | 1.76 | 4.20 | 485.1 |
| **ILD12** | 2.28 | 2048.1 | 7.52 | 1.46 | 2222.8 | 5.18 | 2.18 | 7.52 | 641.3 |
| **ILD13** | 1.66 | 1212.3 | 3.31 | 1.41 | 1380.9 | 3.23 | 1.73 | 3.18 | 543.6 |
| **ILD14** | 1.51 | 1020.9 | 4.53 | 1.53 | 1541.0 | 5.40 | 1.77 | 3.30 | 462.3 |
| **HC01** | 0.37 | 459.2 | 1.25 | 0.40 | 575.4 | 1.54 | n.a | n.a | n.a |
| **HC02** | 0.68 | 651.8 | 2.07 | 0.64 | 800.7 | 2.39 | n.a | n.a | n.a |
| **HC03** | 0.46 | 566.3 | 1.49 | 0.43 | 683.0 | 1.45 | n.a | n.a | n.a |
| **HC04** | n.a | n.a | n.a | n.a | n.a | n.a | n.a | n.a | n.a |

\*The remodeling volumes (R.V.) do not include the basal posterior part of the lung with dependent atelectasis while the lungs volume does. R.V. is lung volume with SUV>0.75 for [<sup>68</sup>Ga]Ga-FAPI-46.

Lung uptake of [^11^C]NES plateaued at 100s post-injection (Fig. 1.A, S5A). In contrast, lung uptake of [^68^Ga]Ga-FAPI-46 reached a stable uptake after 30s but slowly decreased thereafter (Fig.1A). [^11^C]NES uptake in the bone marrow and spleen increased over time (Fig. S5F). Analyses performed on static full-body scans also demonstrated that [^11^C]NES have high uptakes in bone marrow and spleen compare to [^68^Ga]Ga-FAPI-46 (Fig.S7, table.S3).

The ILD patient group exhibited a heterogeneous pattern of tracers uptake (Fig S2,S6). Within the cohort, six patients had subpleural uptake (ILD03, ILD06, ILD11, ILD12, ILD13, and ILD14), six had diffuse uptake (ILD01, ILD04, ILD05, ILD07, ILD09, and ILD10), and two had central uptake (ILD02 and ILD08). For most patients, uptake in the left lung was higher than in the right lung, although not significantly (Fig 2. A, B, table 2,3). Some patients, such as ILD08 and ILD02, showed remarkably higher uptake in the left lung ([^11^C]NES SUV_mean_ in the left whole lung was 2.9 times higher than in the right lung for ILD02, and [^68^Ga]Ga-FAPI-46 uptake was 1.9 times higher in the left whole lung than in the right for ILD08) (Table 2,3). Overall, [^68^Ga]Ga-FAPI-46 SUV_mean_ tended to be higher than [^11^C]NES SUV_mean_ (Fig.2). For example, in ILD08, the SUV_mean_ in the left lung for FAPI was twice as high as the SUV_mean_ for [^11^C]NES, although the R.V. was only 16.7 cm³ higher than the I.V.. Only one patient, ILD03, had a higher [^11^C]NES SUV_mean_ than [^68^Ga]Ga-FAPI-46, with the SUV_mean_ in the I.V. being 1.12 times higher than the R.V., and similar I.V. and R.V. volumes.

**Figure 2.**
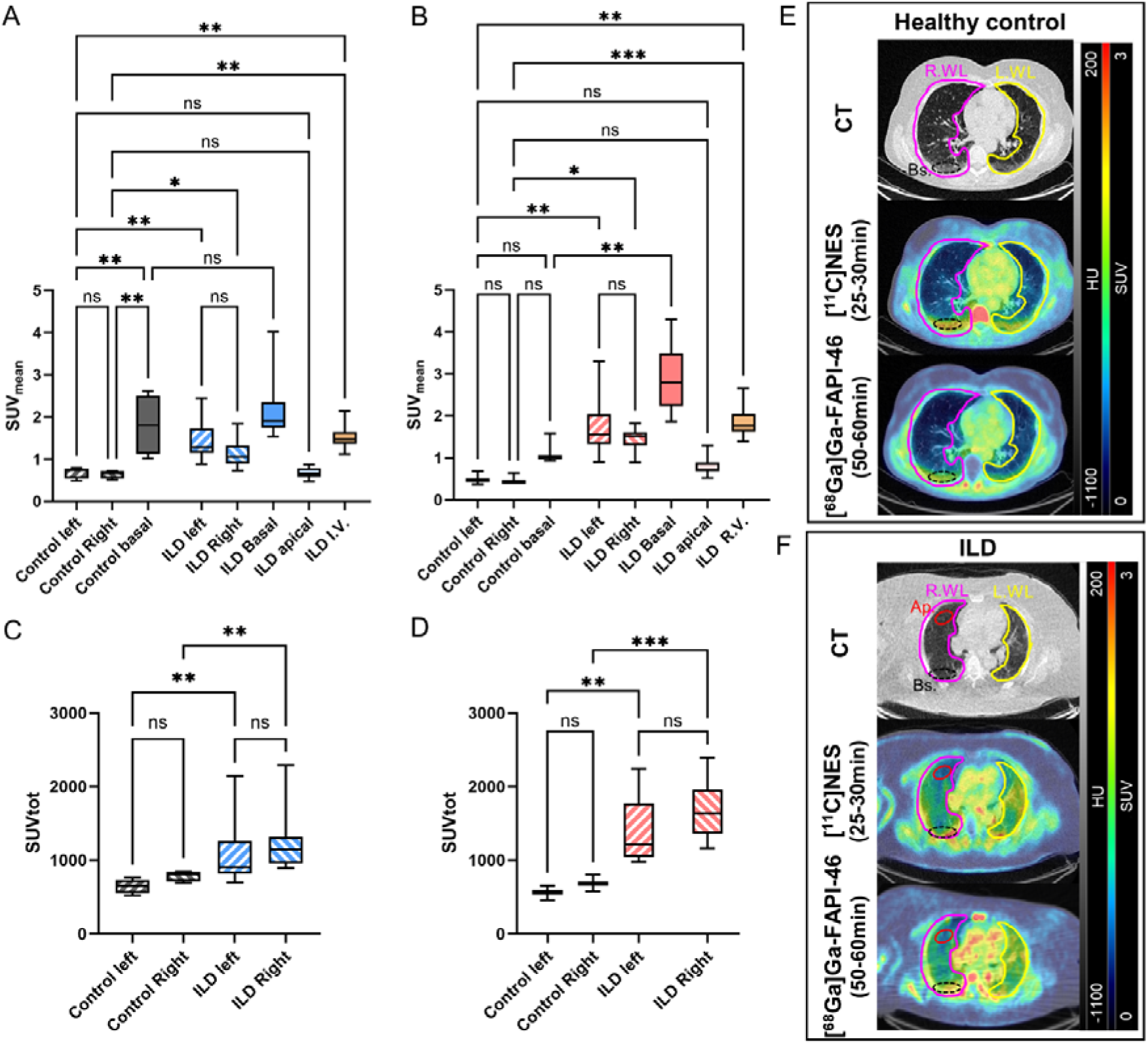
[^11^C]NES **(A,C)** and [^68^Ga]Ga-FAPI-46 **(C,D)** uptake in lungs from ILD patients compared to healthy controls. SUV data obtained in the lung are displayed as Whisker plots. (**A,B)** SUV_mean_ and **(C,D)** SUV_tot_. And representative image of the CT and PET/CT scans in a healthy patient **(E)**, and a patient with UILD and PPF (ILD09) **(F)**. Ap, apical; Bs, basal; HU, Hounsfield unit; I.V.,inflammation volume; R.V., remodeling volume; SUV,standardized uptake value; WL,whole lung.

**Figure 3.**
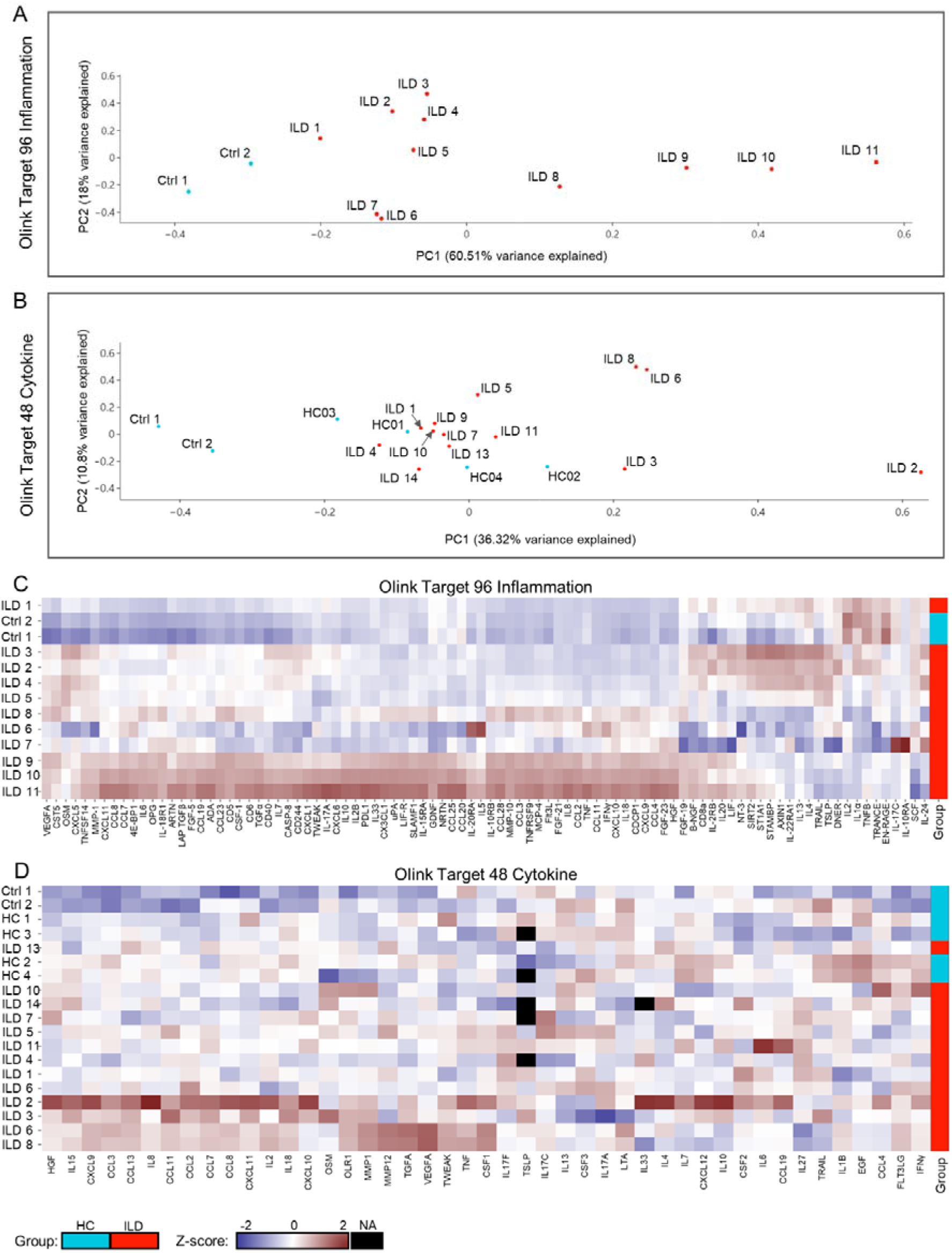
Plasma level of inflammatory biomarkers. (A,B) Heatmap illustrating the plasma levels (expressed as z-scores) of the proteins in ILD patients and healthy controls. **(C,D)** Principal component analysis (PCA). Each point represents a single patient, with patients of similar protein expression profiles positioned next to each other. Results from the Olink Target 96 inflammation panel (ILD n=11, HC n=2) **(A,C)** and Olink Target 48 cytokine (ILD n=13, HC n=6) **(B,D).**

Although tracers uptake were heterogeneous among individuals in the ILD group, SUV_mean_ measurements were higher than those in healthy controls (Fig.2A,B,E,F). Additionally, the mean SUV_mean_ and SUV_tot_ in ILD lungs were significantly higher than in healthy lungs (Fig.2C,D).

### Plasma proteomic analysis

The healthy controls from a previous study (Ctrl1-2) consistently exhibited low levels of inflammatory biomarkers in their plasma across both Olink panels. The cytokines levels in these plasma samples were lower than those observed in the healthy controls from the current study (HC1-4). A key distinction between these two groups is that Ctrl1-2 are younger (in their 20’s), while HC1-4 are of a similar age to the ILD patients (median age (IQR25; IQR75) of the HC group: 63.5 (54.8; 70.0) years, and ILD: 73.5(68.25;78.25) years). The cytokines panel analysis indicated that HC1-4, unlike Ctrl1-2, clustered with many of the ILD patients. The Olink Target 96 Inflammation panel results revealed that the plasma proteomic profile of ILD11, diagnosed with rheumatoid arthritis-associated ILD (RA-ILD), differed from other ILD patients, with overall higher protein levels compared to the other participants. ILD10 and ILD9, both diagnosed with unclassifiable interstitial lung disease (UILD), followed ILD11 in this trend. Among the most differentially expressed proteins between the healthy controls and the ILD groups, those showing the greatest differences included chemokine (C-X-C motif) ligand 5 (CXCL5), CXCL11, CXCL10, chemokine (C-C motif) ligand 8 (CCL8), and interleukin 6 (IL6).

Similarly, the Olink Target 48 Cytokines panel results showed that ILD2, diagnosed with inflammatory UILD, had a plasma proteomic profile distinct from other ILD patients, characterized by elevated cytokine levels, particularly IL8, CXCL12, IL4, IL10, and IL33. Additionally, two of the three patients with nonspecific interstitial pneumonia (NSIP) (ILD6 and ILD8) clustered together and exhibited high plasma cytokine levels, especially vascular endothelial growth factor A (VEGFA), transforming growth factor alpha (TGFA), matrix metallopeptidase 1 (MMP1), MMP12, and colony stimulating factor 1 (CSF1). Among the most differentially expressed proteins between the healthy controls and the ILD groups, the most pronounced differences were observed in CCL7, IL6, CXCL9, MMP1, and oxidized low density lipoprotein receptor 1 (OLR1).

### Correlation between tracers’ uptake, clinical parameters and serum biomarkers

The correlation between PET measures and forced vital capacity (FVC) and six minutes walking distance (6MWD) tended to be negative (Fig.4). For example, I.V. and R.V showed a non-significant negative correlation with FVC (R = -0.52, p = 0.07, and R = -0.47, p = 0.11 respectively). Neutrophil and C-reactive protein (CRP) levels tended to positively correlate with uptake of both tracers. In contrast, hemoglobin levels tended to negatively correlate with I.V. and R.V. volumes, though this was not observed for NES SUV_mean_. No clear correlation could be established between the plasma biomarkers levels measured by relative quantification in the Olink Target 96 Inflammation panel. However, absolute quantification in the Olink Target 48 Cytokine panel revealed that elevated levels of CCL7 and CXCL9 were associated with a higher SUV for both tracers.

**Figure 4.**
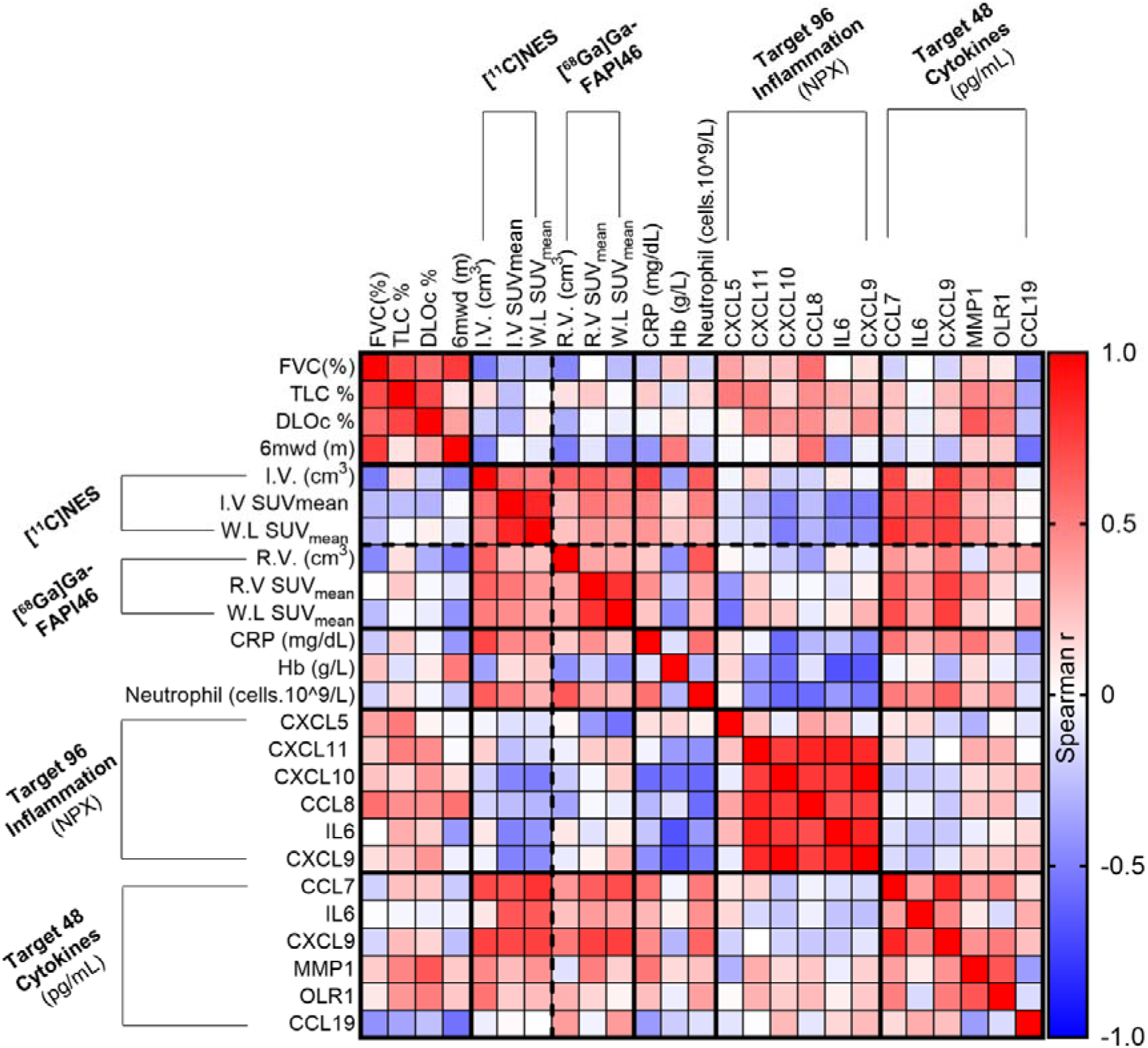
Correlation matrix between PET-derived measures, clinical parameters and plasma proteomic profile. Color intensity indicates the strength of the correlations (Spearman r). The W.L SUV_mean_ represents the SUV_mean_ of the lung with the higher uptake (between left or right lung). DLOc, carbon monoxide diffusing capacity; FVC, forced vital capacity ;Hb, hemoglobin; CRP, C-reactive protein; I.V.,inflammation volume; R.V., remodeling volume; SUV,standardized uptake value; TLC, total lung capacity; WL,whole lung; 6MWD, six minutes walking distance.

## DISCUSSION

In this pilot study, we demonstrated that it is possible to assess neutrophil-mediated inflammation and remodeling activity in the lungs *in vivo* and non-invasively in patients with ILD of varying etiology and severity. Our findings indicate that the tracers accumulate specifically in regions with CT abnormalities, with [^68^Ga]Ga-FAPI-46 showing particularly strong uptake in these areas. In contrast, minimal non-specific signal was observed in healthy lung tissue. These results are consistent with clinical studies that demonstrated the selective visualization of activated fibroblasts in humans using FAPI PET tracers^7–9^ and the detection of neutrophil activity by targeting NE with [^11^C]NES ^10,[manuscript]^.

Lung injury can trigger the recruitment of various immune cells, including neutrophils, which may lead to dysregulated immune responses and subsequent pulmonary fibrosis. Clinical studies have shown that elevated neutrophil levels in the blood and lungs of idiopathic pulmonary fibrosis (IPF) patients are associated with poorer prognoses^11–15^. Neutrophils contribute to fibrosis through the release of NE and neutrophil extracellular traps (NETs), which activate key pro-fibrotic cytokines^16,17^ and promote fibroblast proliferation and differentiation into myofibroblasts^18^. Although single-cell ribonucleic acid sequencing (scRNAseq) analysis of neutrophils in ILD is limited and human ELANE (gene encoding for NE) expression remains unexplored (Fig.S8)^19^, preclinical data show increased ELANE expression in neutrophils from bleomycin-induced fibrotic mice compared to healthy controls^20^. In our study, we demonstrated that [^11^C]NES uptake is significantly higher in the lungs of ILD patients compared to healthy controls, co-localizing with CT signs of inflammation and fibrosis. For example, in patient ILD02, who was diagnosed with UILD and histopathological evidence of inflammation, [^11^C]NES exhibited a markedly high uptake in the left lung, which showed shrinkage and CT signs of fibrosis. [^11^C]NES PET scans may be valuable in guiding treatment decisions, as patients with pronounced neutrophil-mediated inflammation may benefit from anti-inflammatory treatments. Additionally, neutrophil counts in serum positively correlated with [^11^C]NES lung uptake, which also tended to increase as lung function declined (as indicated by low FVC and 6MWD). [^11^C]NES accumulated in organs with high neutrophil concentrations, such as the spleen and bone marrow. [^18^F]FDG, another PET radiotracer used to assess inflammation, has been previously evaluated in IPF and other lung diseases^21^. However, unlike [^11^C]NES, [^18^F]FDG is not specific to a single immune cell population, and will assess all cells with increased glucose consumption leading to increased non-specific [^18^F]FDG uptake in the lungs.

Fibrotic foci are areas within fibrotic regions, characterized by clusters of activated fibroblasts and myofibroblasts which are involved in the excessive deposition of extracellular matrix, playing a pivotal role in the progression of fibrotic diseases^22,23^. FAP expression is selectively induced in these fibrotic foci but is absent in normal lung tissue^6^. scRNAseq analysis of stromal cells has revealed that FAP is expressed in myofibroblasts from IPF patients, with elevated expression in fibroblasts from IPF patients compared to healthy controls, where its expression is minimal (Fig. S8A, B)^19^. The precise role of FAP in lung fibrogenesis remains unclear, some studies suggest it has protective effects through matrix degradation^24^, while others propose that FAP inhibition may offer anti-fibrotic benefits^25^. Nevertheless, FAP remains a promising target for molecular imaging of fibroblast activation and sites of active tissue remodeling. Both preclinical and clinical studies evaluating PET tracers targeting FAP have demonstrated high uptake in fibrotic areas of the lungs, highlighting their potential for monitoring remodeling activity in response to treatment or for the early detection of fibrotic injury^7–9,26,27^. Our findings align with these results, as [^68^Ga]Ga-FAPI-46 showed a high uptake in fibrotic areas, correlating with clinical observation. Dynamic PET scans revealed that [^68^Ga]Ga-FAPI-46 specific uptake in the lung is slowly decreasing over time, in contrast to [^11^C]NES, which is more stable. This suggest that [^11^C]NES bound to NE and is retained, whereas [^68^Ga]Ga-FAPI-46 might have a reversible binding or is being washed out or metabolized over time, this was also reported by Röhrich et al^8^.

In all ILD subjects, a high dorsal uptake of both tracers was observed. A similar pattern, though less pronounced, was also noted in the control group, where a distinguishable dorsal lung uptake was present. This uptake may be attributed to gravity-dependent atelectasis, a condition commonly observed in the posterior lung bases, especially of elderly individuals and patients with lung diseases^28^. To assess the impact of gravity on [^68^Ga]Ga-FAPI-46 uptake, the ILD patient ILD07 was scanned in a prone position, which resulted in decreased dorsal uptake. This finding indicates that the strong uptake observed in the dorsal part of the ILD lungs cannot be solely attributed to fibrosis; gravity-dependent atelectasis must also be considered when analyzing PET/CT scans. Therefore, in our study, the high dorsal uptake was excluded from R.V. and I.V. but was included in the whole lung volume.

We demonstrated that the plasma proteomic profile of ILD patients shows elevated levels of various immune markers compared to controls. For instance, IL6 and IL8, have previously been associated with poor prognosis in IPF patients^33^. Additionally, we identified a correlation between high plasma levels of CCL7 and CXCL9 and increased tracer uptake in the lungs. Previous studies have shown that CCL7, a chemokine involved in neutrophil trafficking to inflamed lungs^34,35^, is expressed at higher levels in biopsies with usual interstitial pneumonia (UIP)^36^, and its plasma level has been associated with disease severity and progression patients with IPF^37^. Similarly, CXCL9, an angiostatic and T-cell chemoattractant, has also been found to be elevated in the plasma or serum of ILD patients and positively correlated with changes in FVC% and TLC%^37–39^. However, for the majority of markers, the proteomic analysis did not reveal a clear correlation with disease severity or tracer uptake in the lungs. Plasma levels of inflammatory markers may be influenced by various comorbidities, such as type 2 diabetes (T2D), obesity and hypertension.

Efforts are being made to enhance the assessment of fibrosis activity to improve patient prognosis. For instance, worsening fibrosis observed on HRCT is associated with an increased risk of mortality. However, the current approach to quantification lacks uniformity and is limited by human error, such as interobserver disagreement. To address these limitations, advancements in HRCT analysis are being explored, including the use of quantitative computer-based imaging analysis methods^40,41^. Despite these improvements, HRCT remains unable to provide functional analysis of inflammation or remodeling activity, which can be achieved through PET imaging.

Molecular imaging with tracers that enable the monitoring of specific immune cells and remodeling activity within the lung, such as those evaluated in this study, holds significant potential. In diseases like ILDs, which represent a highly heterogeneous group of lung disorders, distinguishing between fibrosis-driven and inflammation-driven disease activity could greatly aid in patient stratification, facilitate more effective disease management through personalized treatment, and support the development of anti-fibrotic therapies.

Our study had some limitations. As a single-center pilot study with a small size and heterogenous cohort, including only fourteen patients and four controls, the statistical analyses conducted are exploratory in nature. Further research with larger and more diverse cohorts is necessary to validate and extend these findings. Another limitation of our study is the age disparity between the ILD patients and the control groups in the Olink panel Target 96 Inflammation analysis, with the controls being younger (in their 20’s) than the ILD patients (mean age: 73 years). This age gap introduces a bias, making it difficult to accurately compare the inflammatory proteomic profiles between the two populations.

## CONCLUSION

In this study, we demonstrated that [^11^C]NES and [^68^Ga]Ga-FAPI-46 PET imaging can effectively assess neutrophil-mediated inflammation and tissue remodeling in patients with ILD. The distinct uptake patterns observed in diseased lung regions highlight the potential of PET scans using these tracers as non-invasive tools for diagnosing and monitoring ILD. These findings suggest that [^11^C]NES and [^68^Ga]Ga-FAPI-46 PET imaging could play a crucial role in guiding personalized treatment strategies and advancing the development of targeted anti-fibrotic therapies.

## DISCLOSURE

Olof Eriksson is an employee and a minority shareholder of Antaros Tracer AB. Otherwise, the authors have nothing to disclose.

## KEY POINTS

### QUESTION

Can [^11^C]NES followed by [^68^Ga]Ga-FAPI-46 PET scans effectively visualize and differentiate between inflammation-driven and fibrosis-driven disease activity in ILD patients of varying severity and etiology?

### PERTINENT FINDINGS

Both radiotracers showed higher uptake in diseased lungs compared to healthy controls. The uptake co-localized with CT abnormalities, with [^68^Ga]Ga-FAPI-46 typically demonstrating stronger uptake than [^11^C]NES.

### IMPLICATIONS FOR PATIENT CARE

Non-invasive assessment of neutrophil-mediated inflammation and remodeling in ILD patients could serve as a companion diagnostic tool, guiding personalized treatment decisions and improving patient management. Additionally, this method could monitor disease progression and treatment efficacy, supporting the development of new anti-fibrotic therapies.

## DATA AVAILABILITY

The data used and/or analyzed during the current study are available from the corresponding author upon reasonable request.

## Data Availability

All data produced in the present study are available upon reasonable request to the authors.

## Abbreviations

CCL: chemokine (C-C motif) ligand
CRP: C-reactive protein
CSF: colony stimulating factor
CXCL: chemokine (C-X-C motif) ligand
FAP: fibroblast activation protein
FVC: forced vital capacity
HC: healthy control
HRCT: high-resolution computed tomography
ILDs: interstitial lung diseases
IL: interleukin
IPF: idiopathic pulmonary fibrosis
I.V.: inflammation volume
LAP: latency-associated peptide
MMP: matrix metallopeptidase
NE: neutrophil elastase
NET: neutrophil extracellular trap
NPX: normalized protein expression
NSIP: nonspecific interstitial pneumonia
OLR: oxidized low density lipoprotein receptor
PEA: proximity extension assays
PFTs: pulmonary function tests
RA-ILD: rheumatoid arthritis-associated
ILD R.V.: remodeling volume
scRNAseq: single cell ribonucleic acid sequencing
SUV: standardized uptake value
TGF: transforming growth factor
T2D: type 2 diabetes
UILD: Unclassifiable interstitial lung disease
UIP: usual interstitial pneumonia
VEGFA: vascular endothelial growth factor A
VOI: volume of interest
6MWD: six minutes walking distance

