## Supplementary data for "Investigating Inflammation and Tissue Remodeling in ILD with [^11^C]NES and [^68^Ga]Ga-FAPI-46 PET Imaging"

**PET/CT examination**

Subjects were scanned in the supine position. A low-dose CT scan was performed for anatomical localization and attenuation correction of the PET emission data. Following the CT scan, [^11^C]NES was administered via a controlled bolus injection. The final radiotracer, [^68^Ga]Ga-FAPI-46, was injected at least 2 hours after [^11^C]NES, corresponding to six half-lives of [^11^C]. In both cases, a dynamic PET scan over the chest was initiated at the time of injection.

The dynamic [^11^C]NES scan lasted 30 minutes and was divided into frames of increasing durations: 1 × 10s, 8 × 5s, 4 × 10s, 2 × 15s, 3 × 20s, 4 × 30s, 5 × 60s, and 4 × 300s. The dynamic [^68^Ga]Ga-FAPI-46 scan used the same frame durations, with the addition of 3 × 600s frames. Dynamic images were reconstructed using time-of-flight ordered-subset expectation maximization, including resolution recovery, with 3 iterations, 16 subsets, and a 5-mm Gaussian post-processing filter.

A whole-body low-dose CT and PET scan (3 minutes per bed position) was acquired following the dynamic [^11^C]NES and [^68^Ga]Ga-FAPI-46 PET scans. The static whole-body scan was reconstructed using block-sequential regularized expectation maximization (Q.Clear; GE Healthcare) with a regularization parameter of 500. Participants were allowed to move between the [^11^C]NES whole-body scan and the dynamic [^68^Ga]Ga-FAPI-46 scan.

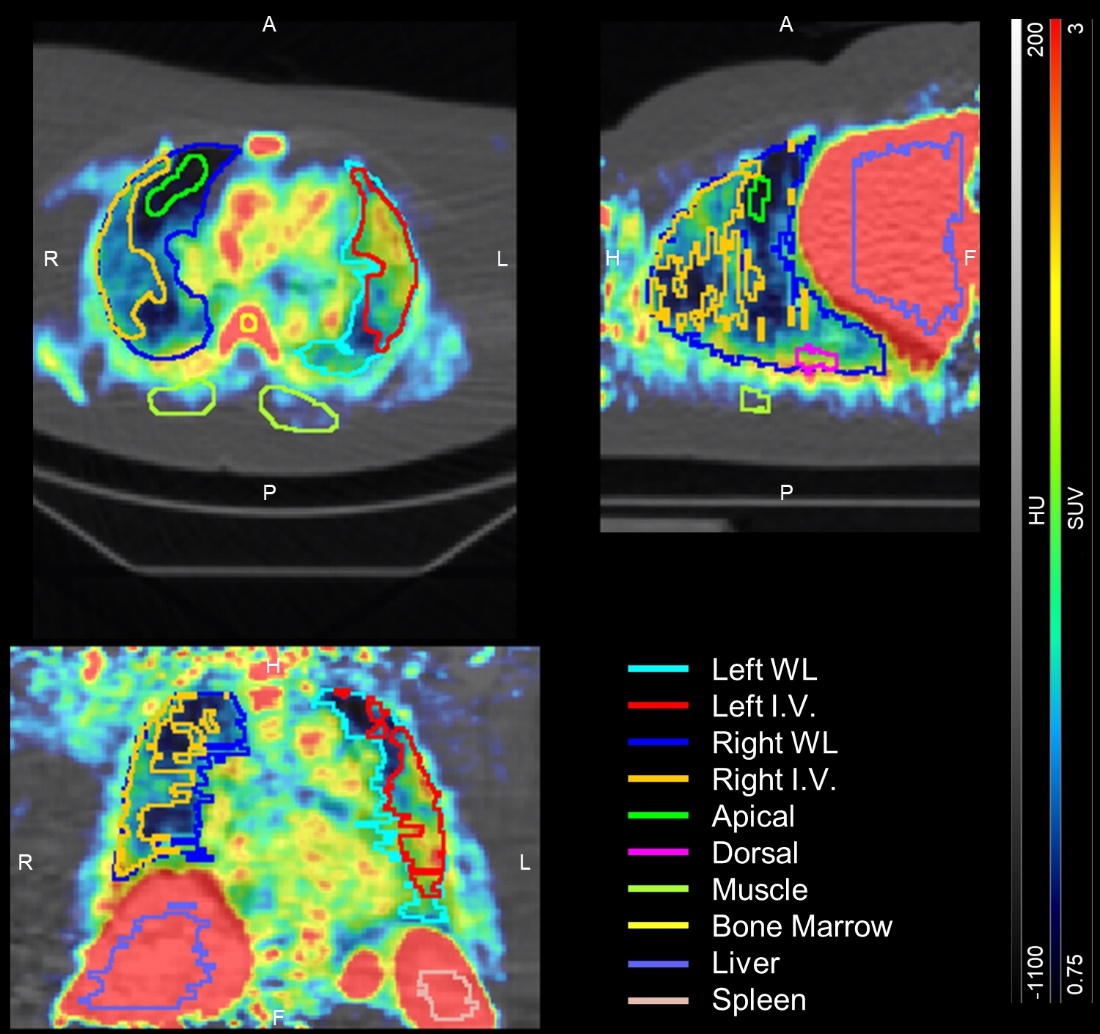

**Figure S1.** Illustration of the methods used to draw the VOIs of the lungs in patient ILD09 [^11^C]NES uptake. The SUV scale was from 0.75 to remove the background signal in the lung. Atelectasis (drawn here as “Dorsal” in pink) and uptake from lobar arteries and veins removed from the inflammation volume VOI. The VOI were drawn on the last frame from the scan, here at 25min. A, apical; F, feet; H, head; I.V. inflammation volume; HU, Hounsfield units ; L, left; P, posterior; R, right; SUV, standardized uptake value ; WL, whole lung.

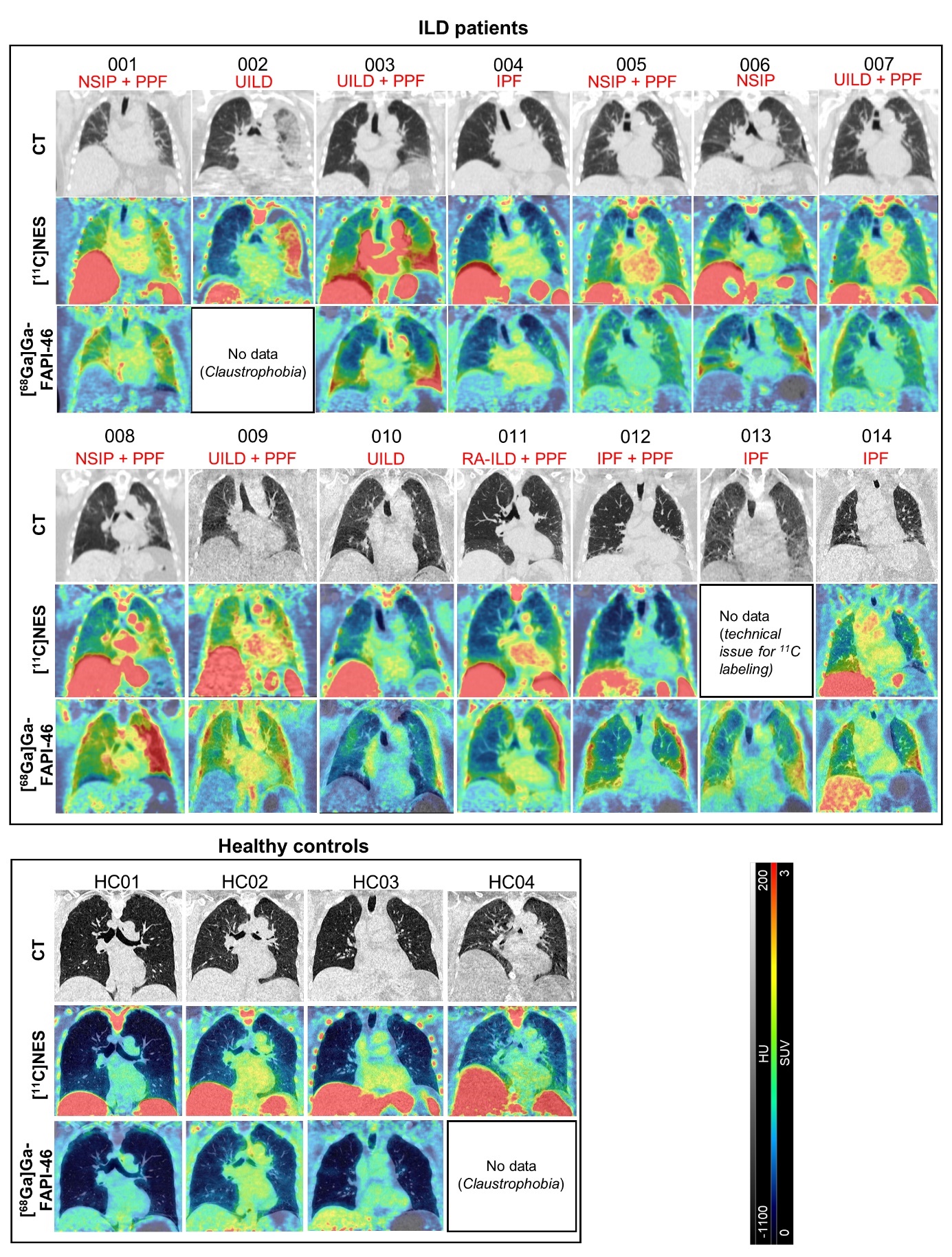

**Figure S2.** Representative axial CT images and PET/CT images of the tracers in all the subjects of the study. The representative chest PET/CT images shown correspond to the last frame of the PET scans. In red is written the diagnostic of each ILD patients. HU, Hounsfield unit; IPF, idiopathic pulmonary fibrosis; NSIP, non-specific interstitial pneumonia; PPF, progressive pulmonary fibrosis; RA-ILD, rheumatoid arthritis-associated ILD; SUV,standardized uptake value; UILD, unclassifiable interstitial lung disease.

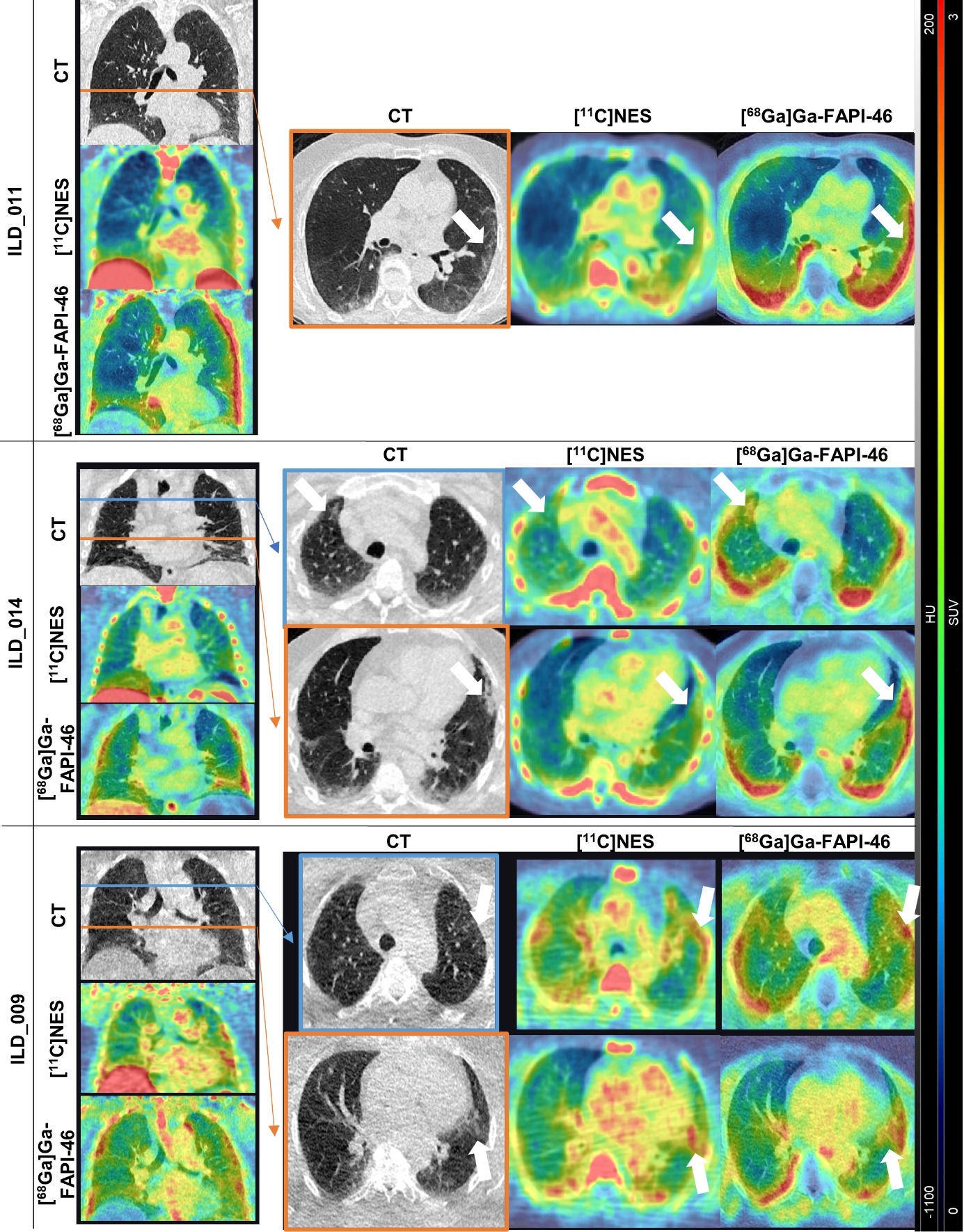
**Figure S3.** Representative coronal and axial CT images and PET/CT images of the tracers’ lung uptake in three different ILD patients: ILD011 (RA-ILD); ILD014 (IPF) and ILD09 (UILD, and PPF). The arrows show CT abnormalities.

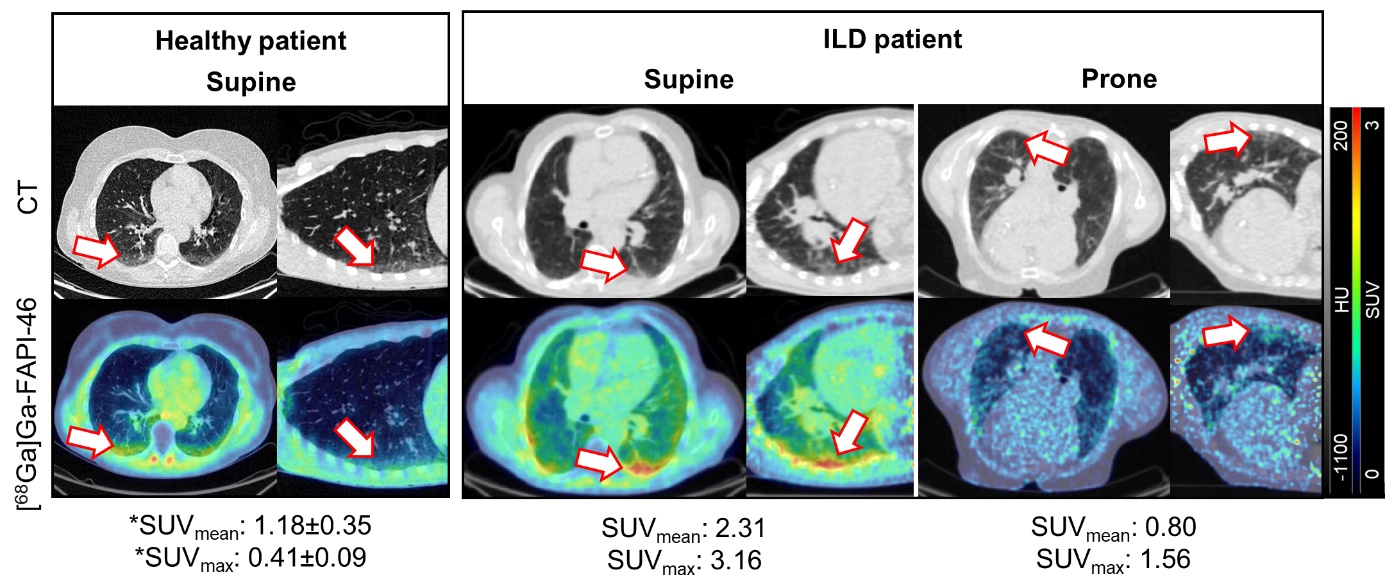
**Figure S4.** Atelectasis and gravitational effect on [^68^Ga]Ga-FAPI-46 dorsal lung uptake. Representative coronal and axial CT images and PET/CT images of the tracers’ lung uptake in a patient with UILD and PPF (ILD07), in supine versus prone position, compared to a healthy control in supine position. The arrows show the dorsal uptake. *The SUV presented are the mean from all the HC (n=3).

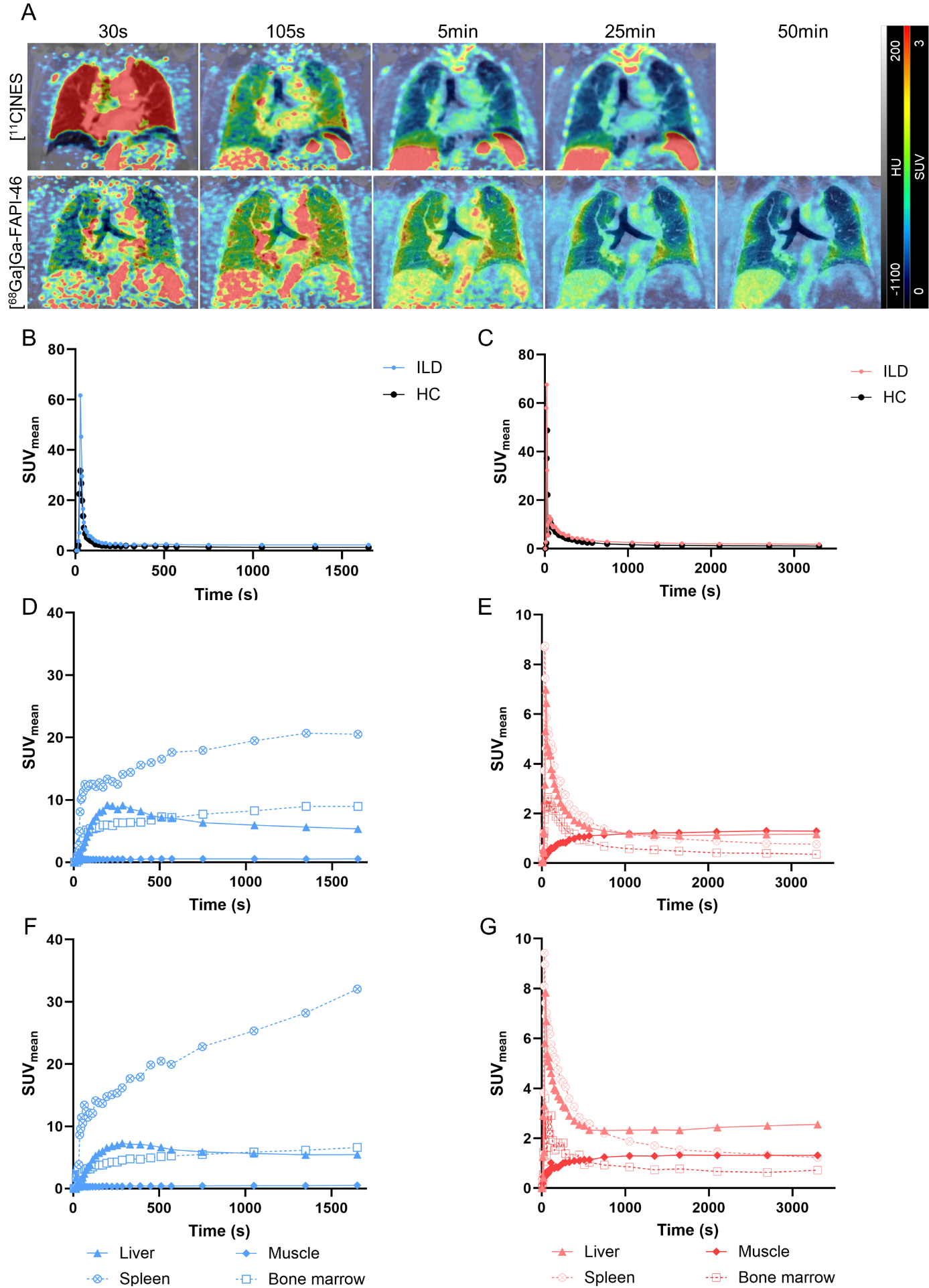

**Figure S5.** Representative axial CT images and PET/CT images of the tracers over the time in a patient with IPF (ILD14) **(A)**. Time–activity curves showing tracers’ aorta uptake in patient ILD14 and a healthy control (HC1) ([^11^C]NES, **(B),** and [^68^Ga]Ga-FAPI-46, **(C)**). Time–activity curves showing liver, spleen, muscle and bone marrow over time of [^11^C]NES **(D,F)** and [^68^Ga]Ga-FAPI-46 **(E,G)**, in ILD14 **(F,G)** and HC1 **(D,E)**  **(C)**.

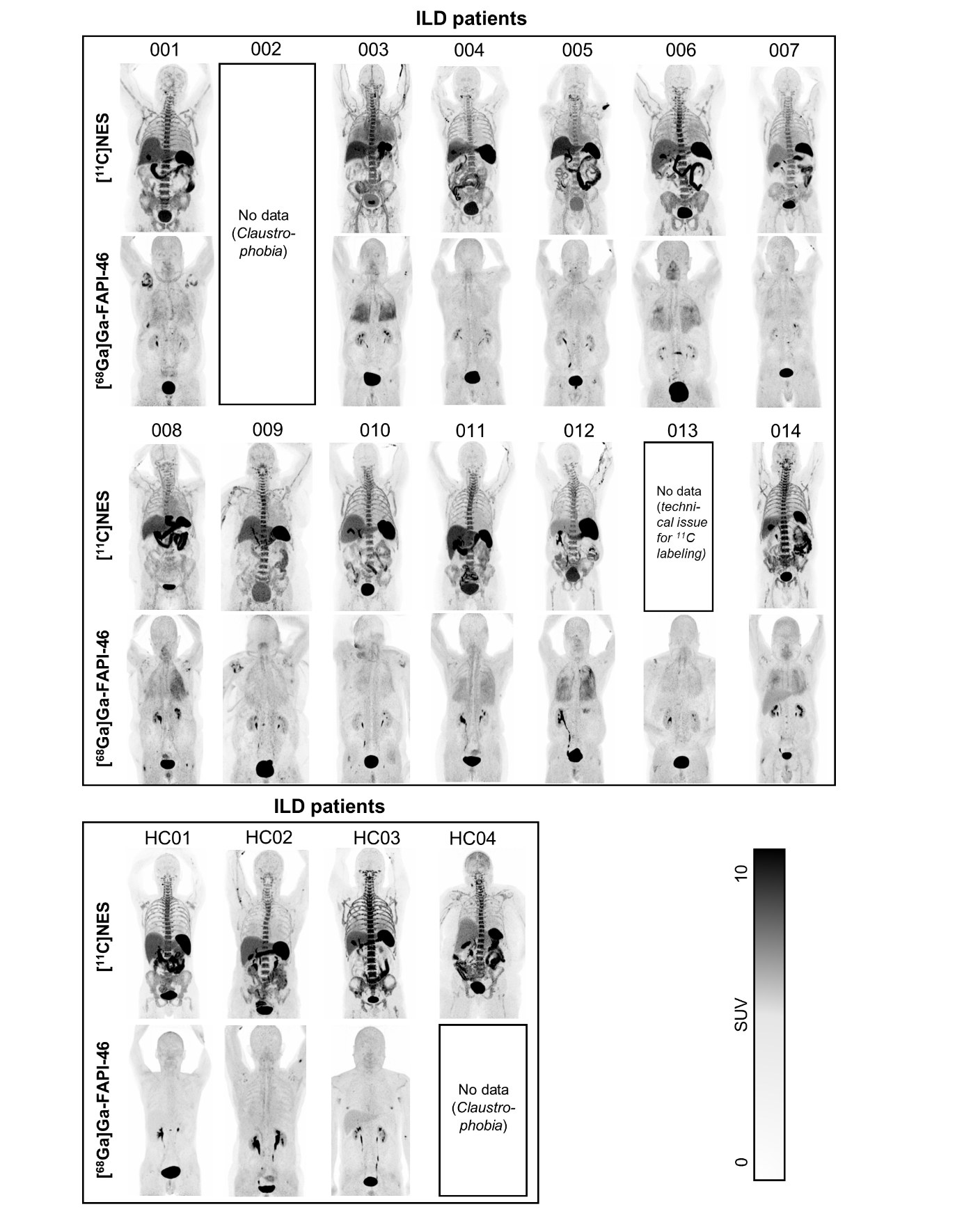
**Figure S6.** Representative maximum-intensity-projection PET images of the tracers full-body uptakes in all the subjects of the study.

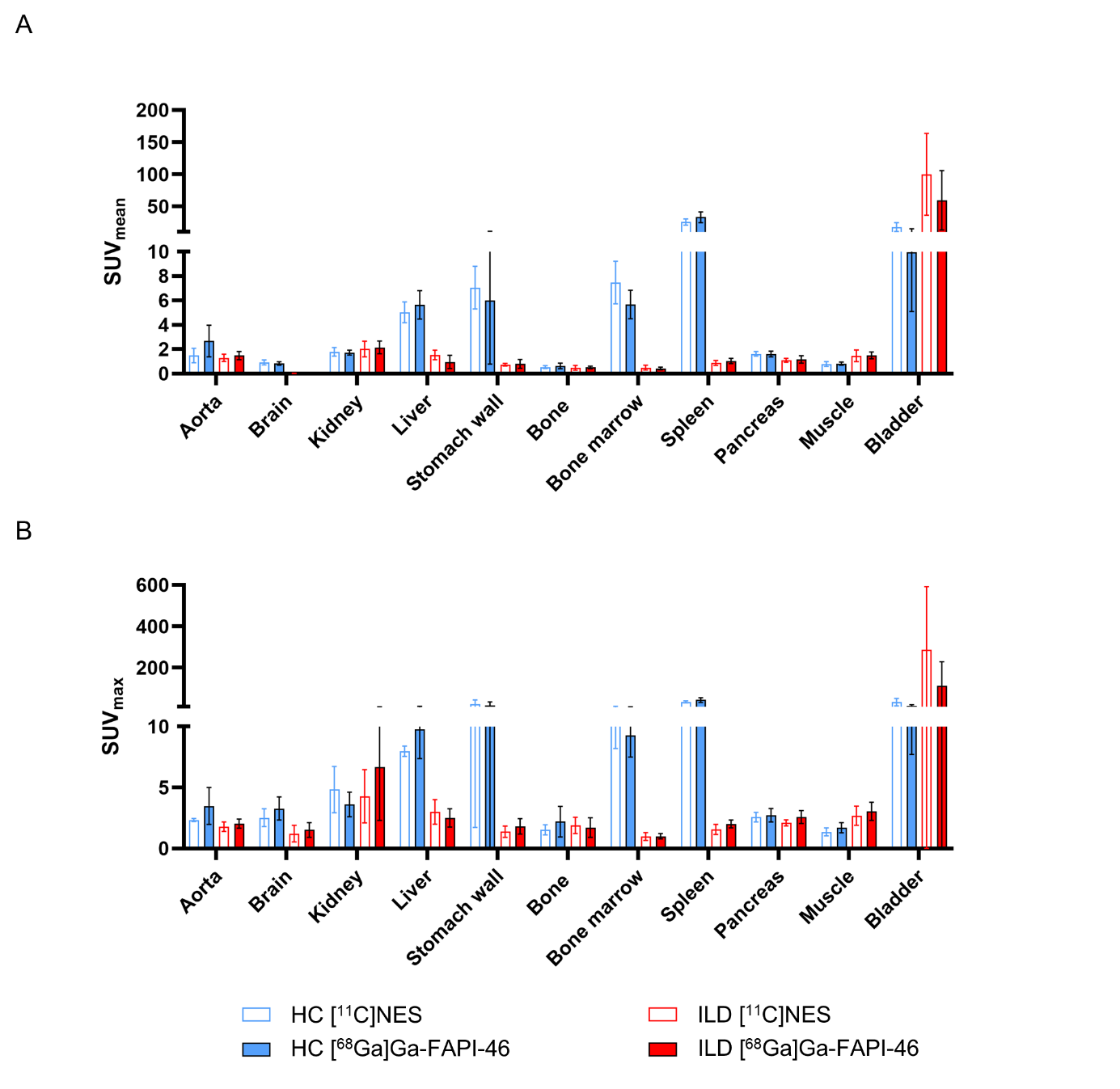

**Figure S7.** Mean SUV_mean_ **(A)** and SUV_max_ **(B)** of [^11^C]NES and [^68^Ga]Ga-FAPI-46 derived from static full body scans in healthy controls (HC) and ILD patients.

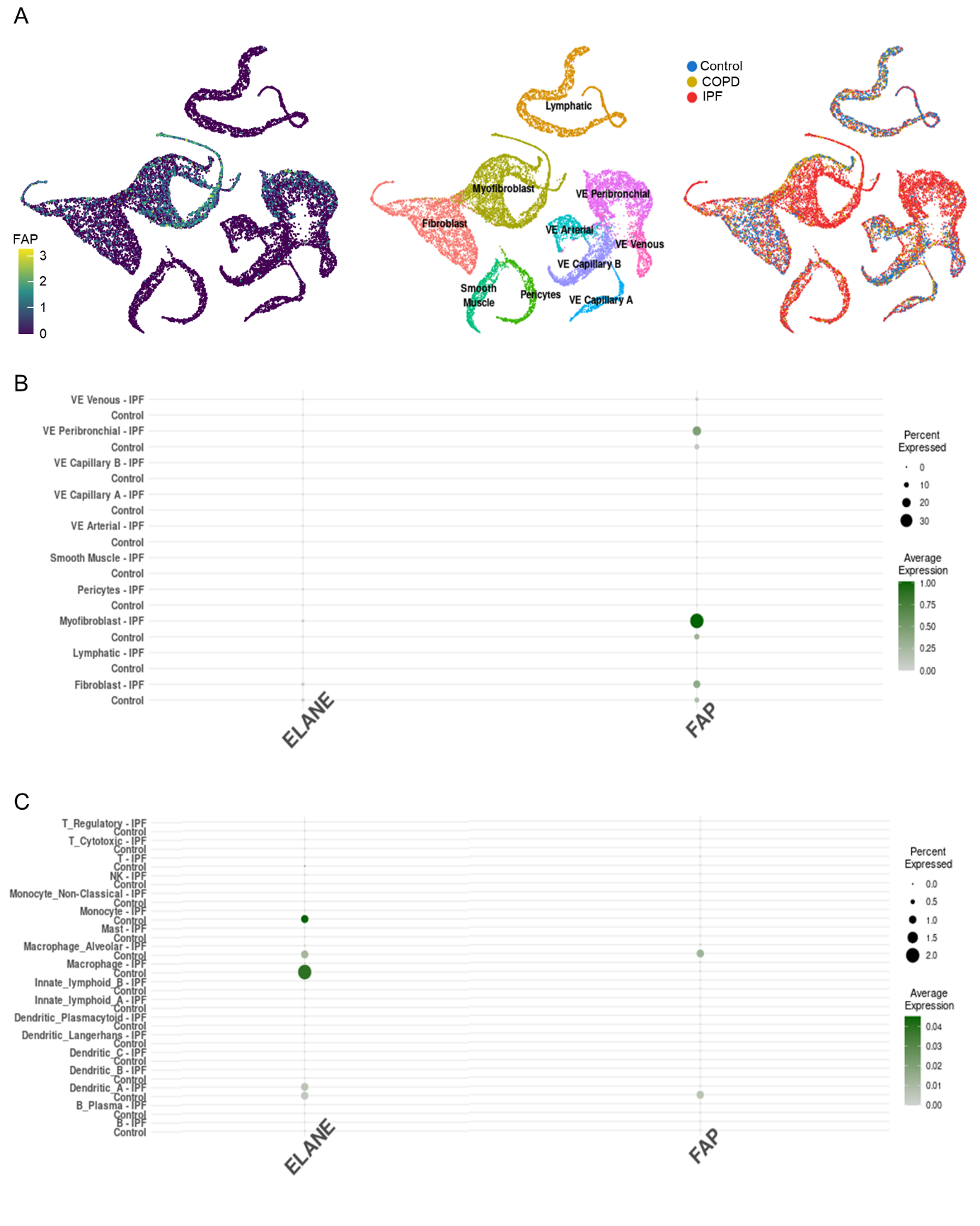
**Figure S8.** (A) UMAP projection on FAP expression in stromal cells from healthy controls, COPD and IPF patients. (B,C) Dot plots of FAP and ELANE gene expression within stromal (B) and immune cells (C). The data is extracted from the IPFcellAtlas.com, from J.McDonough et al. study^19^.

**Table S1.** Clinical characteristics from healthy controls.

| Variable | |  |
| --- | --- | --- |
| Age (Years)* | | 63.5 (54.8; 70.0) |
| Sex | Male | 2 (50%) |
|  | Female | 2 (50%) |
| BMI (kg/m^2^)* | | 23.6 (17.5; 24.7) |
| Smoking status | Former smokers | 0 (0%) |
|  | Never smoked | 4 (100%) |

*Data are presented as median (IQR25; IQR75). BMI, body mass index.

**Table S2.** Patients’ diagnosis, CT patterns and histopathological observations.

| **Patient** | **Diagnosis** | **Progression** | **CT pattern*** | **Histopathological observations** |
| --- | --- | --- | --- | --- |
| ILD01 | NSIP | Yes | Fibrotic NSIP | Normal parenchyma with discrete fibrosis |
| ILD02 | UILD | No | Unclear | Inflammatory disease with secondary discrete fibrosis |
| ILD03 | UILD | Yes | Unspecific fibrotic pattern | n/a |
| ILD04 | IPF | No | Unspecific fibrotic pattern | UIP |
| ILD05 | NSIP | Yes | Fibrotic NSIP | UIP, organizing pneumonia and infection |
| ILD06 | NSIP | No | Inflammatory NSIP with fibrotic components | Discrete fibrosis |
| ILD07 | UILD | Yes | Unspecific fibrotic pattern | n/a |
| ILD08 | NSIP | Yes | Fibrotic NSIP | n/a |
| ILD09 | UILD | Yes | Unspecific fibrotic pattern | n/a |
| ILD10 | UILD | No | Unspecific fibrotic pattern | Increased fibroblasts, mostly normal parenchyma. |
| ILD11 | RA-ILD | Yes | NSIP | n/a |
| ILD12 | IPF | Yes | UIP | n/a |
| ILD13 | IPF | n/a | UIP | n/a |
| ILD14 | IPF | n/a | Probable UIP | n/a |

* Observations were made on a CT scan performed for diagnostic purposes before to the PET examination. BMI, body mass index.IPF, idiopathic pulmonary fibrosis; NSIP, nonspecific interstitial pneumonia; PPF, progressive pulmonary fibrosis; RA-ILD, rheumatoid arthritis-associated interstitial lung disease; UILD, unclassifiable interstitial lung diseases; UIP, usual interstitial pneumonia.

**Table S3:** Static full body PET/CT derived SUVmean measures in all subjects.

|  | **ILD** | | | | | | | | | | | | | **HC** | | | |
| --- | --- | --- | --- | --- | --- | --- | --- | --- | --- | --- | --- | --- | --- | --- | --- | --- | --- |
|  | **01** | **03** | **04** | **05** | **06** | **07** | **08** | **09** | **10** | **11** | **12** | **13** | **14** | **1** | **2** | **3** | **4** |
| **Br** | 2.4 | 6.1 | 2.0 | 3.2 | 2.1 | 2.2 | 4.1 | 2.8 | 1.8 | 2.6 | 1.1 | n/a | 2.0 | 0.6 | 2.0 | 1.8 | 1.6 |
| **Ao** | 0.8 | 0.9 | 0.9 | 0.8 | 0.7 | 0.7 | 0.8 | 1.1 | 0.7 | 0.9 | 0.7 | n/a | 1.0 | 1.2 | 0.9 | 0.7 | 0.9 |
| **Kd** | 1.7 | 2.0 | 1.7 | 1.5 | 1.6 | 1.6 | 1.9 | 1.7 | 1.5 | 1.8 | 1.5 | n/a | 2.1 | 1.5 | 2.3 | 1.8 | 1.6 |
| **Li** | 6.1 | 7.1 | 6.1 | 6.8 | 5.2 | 3.8 | 5.9 | 5.4 | 5.3 | 7.0 | 3.4 | n/a | 5.5 | 5.5 | 5.3 | 5.5 | 3.7 |
| **Sw** | 2.4 | 17.9 | 7.7 | 2.3 | 3.8 | 2.2 | 9.8 | 5.9 | 1.5 | 12.8 | 4.4 | n/a | 1.2 | 5.7 | 7.2 | 9.5 | 5.9 |
| **Bo** | 0.9 | 0.6 | 0.4 | 0.5 | 0.8 | 0.5 | 0.7 | 1.1 | 0.4 | 0.7 | 0.4 | n/a | 0.5 | 0.4 | 0.5 | 0.6 | 0.6 |
| **Bm** | 7.1 | 5.7 | 4.9 | 4.3 | 6.7 | 4.1 | 4.9 | 5.8 | 5.2 | 6.6 | 4.8 | n/a | 7.8 | 6.4 | 5.9 | 9.8 | 7.8 |
| **Sp** | 30.4 | 53.6 | 32.3 | 38.4 | 27.7 | 29.4 | 35.0 | 37.9 | 24.6 | 30.3 | 20.5 | n/a | 36.0 | 24.5 | 30.5 | 28.0 | 19.6 |
| **Pc** | 1.5 | 1.9 | 1.5 | 1.6 | 1.4 | 1.6 | 2.0 | 1.9 | 1.4 | 1.6 | 1.3 | n/a | 1.5 | 1.4 | 1.6 | 1.7 | 1.8 |
| **Mu** | 0.7 | 0.6 | 0.7 | 1.0 | 0.8 | 0.9 | 1.0 | 0.8 | 0.8 | 0.9 | 0.8 | n/a | 0.7 | 0.6 | 1.0 | 0.9 | 0.7 |
| **Bl** | 8.4 | 4.7 | 8.2 | 4.6 | 16.1 | 14.3 | 14.2 | 4.7 | 10.4 | 7.7 | 7.0 | n/a | 19.1 | 14.6 | 16.9 | 27.7 | 12.0 |
| **Br** | 1.3 | 1.5 | 1.8 | 1.8 | 1.2 | 1.1 | 2.0 | 1.8 | 1.0 | 1.7 | 1.1 | 1.2 | 1.8 | 1.0 | 1.6 | 1.3 | n/a |
| **Ao** | 0.0 | 0.0 | 0.0 | 0.0 | 0.0 | 0.0 | 0.0 | 0.0 | 0.0 | 0.0 | 0.0 | 0.0 | 0.1 | 0.0 | 0.0 | 0.0 | n/a |
| **Kd** | 2.0 | 2.7 | 1.9 | 2.3 | 1.7 | 1.8 | 3.3 | 2.4 | 1.8 | 1.8 | 1.7 | 1.8 | 2.8 | 1.3 | 2.6 | 2.1 | n/a |
| **Li** | 0.7 | 0.8 | 0.9 | 1.0 | 0.5 | 0.9 | 1.2 | 0.7 | 0.6 | 0.8 | 0.6 | 1.1 | 2.6 | 1.2 | 1.5 | 1.9 | n/a |
| **Sw** | 0.8 | 1.3 | 1.0 | 0.5 | 0.4 | 0.7 | 1.3 | 0.8 | 0.6 | 1.4 | 0.4 | 0.7 | 0.4 | 0.7 | 0.6 | 0.9 | n/a |
| **Bo** | 0.5 | 0.5 | 0.5 | 0.5 | 0.4 | 0.4 | 0.7 | 0.7 | 0.5 | 0.4 | 0.4 | 0.5 | 0.5 | 0.3 | 0.7 | 0.4 | n/a |
| **Bm** | 0.4 | 0.4 | 0.3 | 0.5 | 0.4 | 0.4 | 0.5 | 0.5 | 0.5 | 0.4 | 0.4 | 0.2 | 0.7 | 0.3 | 0.5 | 0.7 | n/a |
| **Sp** | 0.9 | 1.3 | 1.0 | 1.2 | 0.7 | 1.1 | 1.3 | 1.1 | 0.8 | 1.2 | 1.0 | 0.7 | 1.1 | 0.7 | 1.1 | 0.9 | n/a |
| **Pc** | 1.0 | 1.4 | 1.3 | 1.2 | 0.7 | 0.9 | 1.9 | 1.5 | 1.0 | 1.3 | 1.0 | 0.9 | 1.1 | 1.0 | 1.3 | 1.0 | n/a |
| **Mu** | 1.7 | 1.3 | 1.6 | 1.6 | 1.5 | 1.4 | 1.5 | 1.5 | 1.6 | 2.2 | 1.1 | 1.4 | 1.2 | 1.1 | 2.0 | 1.3 | n/a |
| **Bl** | 62.2 | 49.6 | 42.3 | 47.5 | 41.9 | 55.8 | 30.8 | 34.0 | 48.0 | 55.3 | 37.7 | 57.2 | 210.3 | 36.5 | 14.0 | 99.3 | n/a |

Ao, aorta; Bl, bladder; Bm, bone marrow; Bo, bones; Br, brain; Kd, kidneys; Li, liver; Mu, muscle; Pc, pancreas; Sp, spleen; Sw, stomach wall.

**Table S4.** Summary of the PEA analysis results using the 96 inflammation panel, presenting the NPX means of the most differentially expressed proteins in the serum between healthy controls and the ILD group (the 25 greater NPX difference).

|  | **Control (n=2)** | **ILD (n=11)** | **Difference**  **Control-ILD** |
| --- | --- | --- | --- |
| **CXCL5** | 7.18±0.75 | 10.43±1.37 | -3.25 |
| **CXCL11** | 6.30±0.37 | 9.04±1.21 | -2.74 |
| **CXCL10** | 6.70±0.14 | 9.32±1.80 | -2.62 |
| **MCP-2** | 6.60±0.36 | 9.17±0.96 | -2.57 |
| **IL6** | 1.52±0.28 | 4.04±1.03 | -2.53 |
| **CXCL9** | 5.57±0.10 | 8.10±1.80 | -2.53 |
| **CCL19** | 9.99±0.26 | 11.96±0.98 | -1.97 |
| **MCP-4** | 13.81±0.10 | 15.43±1.20 | -1.62 |
| **EN-RAGE** | 4.51±0.23 | 2.89±0.46 | -1.54 |
| **MCP-3** | 0.68±0.22 | 2.22±0.87 | -1.53 |
| **FGF-21** | 4.26±0.01 | 5.80±1.71 | -1.49 |
| **CDCP1** | 2.55±0.07 | 4.04±1.03 | -1.42 |
| **SIRT2** | 3.03±0.45 | 4.45±0.90 | -1.35 |
| **TNFSF14** | 3.55±0.26 | 4.90±0.49 | -1.34 |
| **MMP-10** | 7.16±0.03 | 8.50±1.03 | -1.29 |
| **LAP TGF-beta-1** | 5.63±0.19 | 6.92±0.50 | -1.26 |
| **HGF** | 7.68±0.07 | 8.94±0.84 | -1.22 |
| **MMP-1** | 12.88±0.24 | 14.10±0.81 | -1.19 |
| **OPG** | 9.50±0.13 | 10.70±0.42 | -1.18 |
| **IL8** | 5.13±0.02 | 6.31±1.21 | -1.18 |
| **IFN-gamma** | 6.22±0.01 | 7.39±0.96 | -1.17 |
| **CCL3** | 4.95±0.06 | 6.13±0.83 | -1.14 |
| **4E-BP1** | 7.96±0.13 | 9.10±0.50 | -1.06 |
| **STAMBP** | 4.61±0.35 | 5.67±0.21 | -1.0596 |
| **CXCL1** | 7.094±0.22 | 8.11±0.31 | -1.0125 |

**Table S5.** Summary of the PEA analysis results using the 48 cytokine panel, presenting the concentrations (median (IQR25;IQR75) of the most differentially expressed proteins in the serum between healthy controls and the ILD group (the 20 greater Log conc. difference).

|  | **Control (n=6)**  **µg/L** | **ILD (n=13)**  **µg/L** | **Log concentration difference**  **Control-ILD** |
| --- | --- | --- | --- |
| **CCL7** | 0.29 (0.28;0.50) | 0.97 (0.57;1.26) | -1.08 |
| **IL6** | 1.88 (1.52;4.47) | 4.41 (3.38;8.59) | -1.05 |
| **CXCL9** | 94.28 (46.24;120.64) | 136.90 (102.09;271.94) | -0.88 |
| **MMP1** | 418.62 (363.39;699.24) | 872.56 (682.33;2043.01) | -0.84 |
| **OLR1** | 47.65 (35.18;56.27) | 83.40 (65.80;118.09) | -0.69 |
| **CCL19** | 102.90 (65.57;163.40) | 172.41 (131.55;227.56) | -0.68 |
| **CXCL10** | 169.92 (74.56;194.45) | 166.99 (125.43;350.62) | -0.64 |
| **CCL13** | 95.16 (52.98;129.89) | 115.84 (89.78;227.26) | -0.58 |
| **IL17F** | 0.29 (0.17;0.78) | 0.82 (0.37;1.26) | -0.55 |
| **MMP12** | 162.88 (155.50;244.85) | 242.92 (166.20;627.21) | -0.54 |
| **OSM** | 2.42 (2.21;3.11) | 3.08 (2.55;5.07) | -0.49 |
| **CCL11** | 79.05 (71.94;94.98) | 109.18 (93.65;155.73) | -0.49 |
| **CCL8** | 25.65 (23.00;38.04) | 29.76 (25.91;43.43) | -0.45 |
| **CCL2** | 303.96 (267.88;411.19) | 477.68 (416.31;569.11) | -0.45 |
| **HGF** | 263.38 (214.46;391.80) | 423.87 (288.14;486.31) | -0.44 |
| **IFNG** | 0.17 (0.12;0.43) | 0.25 (0.20;0.37) | -0.41 |
| **IL2** | 0.01 (0.01;0.02) | 0.01 (0.01;0.02) | -0.41 |
| **TSLP** | 0.05 (0.03;0.09) | 0.03 (0.05;0.09) | -0.38 |
| **CXCL8** | 6.58 (5.16;9.09) | 8.83 (6.77;10.36) | -0.37 |
| **CCL3** | 5.93 (3.40;7.53) | 7.02 (5.75;8.80) | -0.35 |
